# The GI-specific Avoidance Scale (GIAS): Development, psychometric validation, and incremental power of a new questionnaire

**DOI:** 10.64898/2026.02.23.26346871

**Authors:** Inês A. Trindade, Ana Pereira, Bruna Veloso, Tom van Gils, Sanna Nybacka

## Abstract

**Background and Aims:** Avoidance of symptom-related situations is common in chronic gastrointestinal (GI) conditions, contributing to greater symptom severity, psychological distress, and reduced quality of life. However, no validated measure exists to comprehensively assess GI-specific avoidance. We developed and validated the GI-specific Avoidance Scale (GIAS), a self-report instrument measuring behavioral and cognitive avoidance specific to GI symptoms.

**Methods:** Following literature review and multidisciplinary input, an initial pool of 58 items was generated and refined through expert and patient ratings, yielding 37 items. A sample of 102 adults (mean age 40.8 years) with medically diagnosed GI conditions completed the GIAS and validated measures of avoidance, psychological flexibility, illness shame, GI symptoms, distress, and quality of life. Exploratory factor analysis was used to determine factor structure. Internal consistency, convergent validity, incremental validity, and mediation analyses were conducted.

**Results:** Factor analysis supported a 20-item, three-factor solution: General Avoidance, Food Avoidance, and Intimacy/Body Exposure Avoidance. Internal consistency was excellent for the total scale (α = .94) and good-to-excellent for subscales (α = .82–.94). GIAS scores correlated positively with illness shame, GI symptoms, and distress, and negatively with psychological flexibility, self-compassion, and quality of life. GIAS showed incremental validity over a general illness avoidance measure (IBAS) in predicting GI symptoms and anxiety. Moreover, mediation models suggested that GI-specific avoidance partially mediates bidirectional associations between GI symptoms and psychological distress.

**Conclusions:** The GIAS is a novel, psychometrically robust, and multidimensional self-report questionnaire of GI-specific avoidance. It holds potential for clinical assessment, treatment planning, and evaluation of intervention mechanisms in GI populations.

## Introduction

Experiential avoidance is a broad and common psychological process that serves the basic evolutionary function of protecting the organism from perceived threats or discomfort (1, 2). This process, referred to in this study as avoidance henceforward, refers to the unwillingness to be in contact with distressing internal experiences such as sensations, thoughts, or emotions, and can take form, for example, as thought suppression, attentional control (e.g., hypervigilance), and behavioral withdrawal or inhibition (Hayes, 1996 #4). While avoidance is often useful in the short term by reducing distress or preventing harm (with a few exceptions, such as thought suppression, which usually produces a paradoxical rebound effect; (3)), excessive or rigid avoidance in response to internal experiences often impairs physical health. This process is therefore considered one of the key transdiagnostic mechanisms underlying psychopathology and functional impairment (1, 3).

Avoidance is frequently observed in individuals with chronic gastrointestinal (GI) conditions (4, 5). It is commonly used as a strategy to prevent anticipated GI symptoms, avoid feeling embarrassment or shame, or reduce fear, for instance, through food avoidance/restriction, limitation of social or professional engagements, or always planning routes with toilet access in mind (6–9). These strategies are often operantly reinforced by the immediate relief of experiencing less anxiety or symptom anticipation, but usually lead to reduced social functioning, food-related anxiety, excessively restrictive diets, increased GI symptom severity, and healthcare overutilisation in the long-term (4, 7, 10–12).

In the context of psychological treatment, avoidance is both a key mechanism and a treatment target. Exposure-based interventions, such as CBT or internet-delivered exposure therapy protocols (13, 14), and acceptance-based therapies, such as Acceptance and Commitment Therapy (ACT; (15)) explicitly aim to reduce maladaptive avoidance by promoting repeated, safe contact with feared or avoided stimuli (e.g., food, symptoms, difficult emotions). This weakens the conditioned fear response and promotes tolerance of distressing internal experiences, which ultimately increases behavioural flexibility and clinical improvement (e.g., in irritable bowel syndrome (IBS; (10, 16)).

However, despite the relevance of this process, the field lacks a GI-specific assessment tool that captures the full, and specific, scope of avoidance in these populations. A commonly used instrument in this context is the Visceral Sensitivity Index (VSI; (17)), but it only partially addresses avoidance, focusing either on anxiety about visceral sensations or on generic avoidance unrelated to GI-specific contexts (e.g., “I often worry about the possibility of not having a bathroom available”). Other scales, such as the Illness Behavior Avoidance Scale (IBAS; (18)) measure avoidance tendencies more broadly and are not designed to capture content- or context-specific behaviours relevant to GI symptoms. Thus, these tools may overlook key experiential and behavioural dimensions of GI-related avoidance.

The lack of comprehensive, validated tools limits our ability to accurately identify avoidance patterns in response to experiences related to GI conditions, to adequately evaluate the mechanisms of change of behavioural interventions, and to tailor clinical care. To address this gap, this study aimed to develop and validate the GI-specific Avoidance Scale (GIAS), the first self-report questionnaire measuring behavioural and attitudinal avoidance specific to GI symptoms.

## Methods

### Procedures

This study was approved by the Ethics and Deontology Committee of the Faculty of Psychology and Educational Sciences at the University of Coimbra, Portugal (CEDI/FPCEUC: 89/7), and was conducted in accordance with the Declaration of Helsinki. All participants gave written informed consent.

Data collection took place in Portugal, with participants recruited through targeted outreach via social media platforms and national patient associations. Eligibility criteria were: a) age ≥18 years; b) self-reported medical diagnosis of a GI condition; c) fluency in Portuguese; d) not being pregnant.

Data were collected via self-report questionnaires hosted on LimeSurvey (v.6). To ensure data quality, three attention-check questions were embedded throughout the survey. Of the 255 individuals who accessed it, 104 completed the entire protocol. Two of these failed ≥ 2 das 3 of the attention questions and were excluded. Therefore, a sample of 102 participants was included in the analyses. The full description of this study’s Procedures can be found as Supplementary Material 1.

### Scale Development

Drawing on existing empirical literature in GI conditions (5, 10, 19), clinical theory (e.g., ACT model of psychological flexibility; (15)), and the team’s multidisciplinary expertise (including psychologists, two gastroenterology consultants, one gastroenterology resident, an one dietician), GIAS was designed to capture core domains of GI-related avoidance. These include behavioural, cognitive, and interpersonal avoidance patterns frequently observed in GI populations (e.g., planning daily routines around symptoms, suppressing distressing thoughts or emotions, restricting foods out of fear of triggering symptoms, avoiding physical closeness due to symptom-related shame or body image concerns). The chosen response scale was a 6-point Likert scale, ranging from 0: Strongly disagree to 5: Strongly agree.

An initial pool of 58 items was generated based on existing literature, clinical experience, and team consensus. These items were independently rated for content relevance by 3 external experts (2 psychologists and 1 gastroenterologist) and 1 patient representative, using a 3-point Likert scale (1 = not relevant, 2 = somewhat relevant, 3 = highly relevant). Items were retained if achieving a mean score (M) of 2.5 or higher. Specifically, 9 items were unanimously rated as highly relevant (M = 3.0), and 19 items received an average of 2.75, with three of four raters deeming them highly relevant and one expressing neutrality. An additional 5 items, with an average score of 2.5, were retained based on majority agreement (n = 3), with only one rater deeming them non-relevant. One new item, proposed by an expert, was incorporated into the final pool. In addition, four items were revised for clarity and precision. The resulting preliminary version of the GIAS comprised 37 items.

### Measures

#### Sociodemographic and clinical data

Participants self-reported sociodemographic data such as age, sex, gender identity, marital status, and education level. Furthermore, clinical variables were also self-reported, and included GI diagnosis, current symptoms, and ongoing treatments or medications.

#### Illness behavior avoidance (IBAS)

The IBAS (18) is a 7-item self-report scale designed to assess illness-related experiential avoidance. Items reflect strategies such as avoiding thoughts about the illness, steering clear of symptom-triggering situations, attempting to control symptoms, and concealing symptoms. Responses are rated on a 5-point Likert scale ranging from 0 (“Never”) to 4 (“Always”).

#### Psychological flexibility (CompACT-8)

The CompACT-8 (20–22) is an 8-item version of the original 23-item Comprehensive Assessment of ACT Processes, validated for Portuguese illness groups. It measures psychological flexibility through two core subscales: Behavioral Awareness and Valued Action. Participants rate each item on a 7-point Likert scale from 0 (“Never true”) to 6 (“Always true”).

#### Illness shame (CISS)

The CISS (23) is a 7-item scale that evaluates the level of shame experienced in relation to living with a chronic illness or its symptoms. Items are rated on a 5-point Likert scale from 0 (“Never true”) to 4 (“Always true”).

#### Self-compassion (SCS-SF)

The SCS-SF (24, 25) is a 12-item questionnaire assessing the components of self-compassion. Items are rated on a Likert scale, from 1 (“Almost Always”) to 5 (“Almost Never”).

#### GI symptoms

The frequency of GI symptoms over the past month was assessed using a self-report inventory adapted from previous research (26). Items evaluate the occurrence of common GI symptoms, including abdominal pain, bloating, loose stools, urgency, excessive gas, tenesmus, frequent bowel movements, nausea, and difficulty sleeping due to GI discomfort. Respondents rate each symptom on a 7-point Likert scale ranging from 0 (“Never”) to 6 (“Always”),.

#### Depressive symptoms and anxiety (DASS-21)

The DASS-21 (27) assesses symptoms of depression, anxiety, and stress. Each subscale contains 7 items rated on a 4-point Likert scale from 0 (“Did not apply to me at all”) to 3 (“Applied to me very much or most of the time”). In this study, only the depression and anxiety subscales were used.

#### Quality of Life (EUROHIS-QOL-8)

The EUROHIS-QOL-8 (28) is a questionnaire adapted from the WHOQOL-BREF, with 8 items, each scored from 1 to 5 (“Very dissatisfied” / “Not at all”/ “Very poor” to “Very satisfied”/ “Completely” / “Very Good”). It assesses overall quality of life across four core domains: physical health, psychological well-being, social relationships, and environmental conditions.

A full description of these measures can be found as Supplementary Material 2. To test the internal consistency of the included measures in this study’s sample, Cronbach’s alphas were calculated for the IBAS, CompACT-8, CISS, SCS-SF, DASS-21, and EUROHIS-QOL-8. All alphas revealed good to excellent internal consistencies (all ≥ 0.80).

### Data analysis

All analyses were conducted using IBM SPSS Statistics (Version 29). Exploratory factor analysis (EFA) with principal axis factoring and oblimin rotation was performed to examine the structure of the GIAS. The number of factors was determined based on eigenvalues greater than 1, scree plot inspection, and theoretical interpretability. Items with low communalities (< .35; (29)), factor loadings below .40 (30), or substantial cross-loadings were removed (29). Internal consistency of the final scale and subscales was assessed using Cronbach’s alpha, with values ≥ .70 considered acceptable, ≥ .80 good, and ≥ .90 excellent (31)

Pearson correlations were used to examine GIAS’s relationships with the other studied questionnaires. Hierarchical regression analyses were conducted to assess the incremental validity of the GIAS above and beyond IBAS in predicting GI symptoms, psychological distress (calculated by summing DASS-21’s anxiety and depressive symptoms subscales), and quality of life. Finally, mediation analyses were performed using PROCESS macro (Model 4; (32)) with 5,000 bootstrap resamples to test whether GI-specific avoidance mediated the associations between GI symptoms and psychological distress (anxiety + depressive symptoms). Confidence intervals not including zero were interpreted as demonstrating significant indirect effects.

## Results

### Participants

The final sample (Table 1) included 102 Portuguese individuals with ages between 18 and 78 years (M = 40.82). Most participants were female and identified as a woman. Participants reported a range of GI diagnoses: most having IBS or inflammatory bowel disease (IBD), and 3.9% reported other GI conditions (e.g., gastric reflux).

**Table 1.** Sociodemographic and clinical data of participants (*N* = 102).

|  |  |  |
| --- | --- | --- |
| **Age, *M*, *SD*** |  | 40.82, 12.65 |
| **Sex, %, *n*** |  |  |
| Male |  | 21.6, 22 |
| Female |  | 78.4, 80 |
| **Gender, %, *n*** |  |  |
| Cisgender woman |  | 63.7, 65 |
| Cisgender man |  | 18.6, 19 |
| Transgender man |  | 1, 1 |
| Not reported |  | 16.7, 17 |
| **Education level, %, *n*** |  |  |
| Up to secondary school |  | 32.4, 33 |
| University education |  | 67.6, 69 |
| **Marital status, %, *n*** |  |  |
| Single |  | 39.2, 40 |
| Married or in a cohabitating |  | 55.9, 57 |
| Separated or divorced |  | 4.9, 5 |
| **Diagnosis, %, *n*** |  |  |
| Irritable Bowel Syndrome |  | 42.2, 43 |
| Crohn's Disease |  | 18.6, 19 |
| Ulcerative Colitis |  | 20.6, 21 |
| Celiac Disease |  | 14.7, 15 |
| Other |  | 3.9, 4 |

### Exploratory Factor Analysis

An EFA was conducted using principal axis factoring with oblimin rotation to examine the structure of the GIAS (37 items; Supplementary Material 3). The suitability of the data for factor analysis was confirmed by the Kaiser–Meyer–Olkin (KMO) measure of sampling adequacy, which was .882, and Bartlett’s test of sphericity, which was significant, χ²(666) = 2743.67, p < .001. Eight factors with eigenvalues greater than 1.0 were identified, and upon inspection of item factor loadings, three factors were considered to be pertinent.

Therefore, a second principal axis factoring analysis with oblimin rotation was performed, forcing the extraction of three factors. The first factor explained 37.64% of the variance of the scale, the second factor 6.16%, and the third factor 5.36%. Ten items (1, 3, 4, 6, 7, 8, 11, 13, 27, and 37) were removed for loading on more than one factor and/or having a factor loading below .40.

A third analysis was then performed without these items, revealing three items (5, 16, and 19) with communalities below .35, which were removed. Moreover, because several items showed high correlations with other items assessing the same or very similar constructs, further items were removed. Decisions to remove one of the items of a highly related pair were based on communalities (the item removed had the lowest communality) and/or correlations with other items (the item showing more high correlations with other items was removed). This led to the removal of four items (10, 23, 26, and 28).

The final analysis showed 20 items with factor loadings from .47 to .96, clearly belonging to one of three factors (Table 2; original Portuguese version in Supplementary Material 4). The first factor, with 11 items (12, 14 ,15, 17, 21, 22, 25, 33, 34, 35, and 36), seemed to portray a general avoidance theme, reflecting multiple forms of psychological and behavioural avoidance, often discussed in the context of cognitive avoidance (e.g., “I try to stay busy to keep thoughts or feelings about my GI symptoms from coming”), control or safety-seeking (e.g., “I plan to go out only when I know there is a toilet nearby”), self-concealment (e.g., “I don’t leave my home if I think my GI symptoms will be noticed by others”), and social withdrawal (e.g., “I tend to move away from friends and family when my GI symptoms get worse”).

**Table 2.** The final 20 items and three subscales of the GIAS.

| Item | Subscale |
| --- | --- |
| 12. I try to keep myself busy to avoid having negative thoughts or feelings about my GI symptoms. | General Avoidance |
| 14. When I have GI symptoms, I usually change my plans for that day. |  |
| 15. I avoid meeting new people because I'm afraid of being judged for my GI condition. |  |
| 17. I distance myself from friends and family when my GI symptoms worsen. |  |
| 21. I don't make long-term plans due to the unpredictability of my GI condition. |  |
| 22. I avoid leaving home when I believe other will notice my GI symptoms. |  |
| 25. I only make plans to go out when I know there will be a toilet nearby. |  |
| 33. I avoid using public transports due to my GI symptoms. |  |
| 34. I make an effort to avoid thinking about my fear of having a new flare related to my GI condition. |  |
| 35. I try not to be too far from a toilet to avoid feeling anxious. |  |
| 36. I struggle attending social events because I'm afraid of having GI symptoms in public. |  |
| 2. I rarely try new foods because I'm afraid of having GI symptoms. | Food Avoidance |
| 9. I avoid drinks or foods that I enjoy because I fear they will trigger GI symptoms. |  |
| 20. I avoid going to restaurants because of my GI condition. |  |
| 30. I avoid certain foods because I fear they will worsen my GI symptoms. |  |
| 31. The way I feel I have to eat due to my GI condition has limited my lifestyle. |  |
| 18. I don't have sex as much as I would like to because I don't want to show signs of my GI condition. | Intimacy and Body Exposure Avoidance |
| 24. I avoid wearing clothes I like, especially tight ones, due to my GI symptoms (for example, feeling bloated). |  |
| 29. I avoid being naked in front of others because I don't want to show signs of my GI condition. |  |
| 32. I avoid having sex because I'm afraid of faecal incontinence or passing gas. |  |

The second factor, with 5 items (2, 9, 20, 30, and 31) specifically included items related to eating and diet, and therefore was named Food Avoidance. Lastly, the third factor portrayed sexual avoidance (“I avoid having sex because I fear faecal incontinence or gas) and physical appearance-related avoidance (“I avoid being naked in front of others because I don’t want to show signs of my GI condition”), and therefore was named Intimacy and Body Exposure Avoidance.

### Reliability Analysis

The internal consistency of the final 20-item scale was excellent, with a Cronbach’s alpha of .94 for the total scale. The General Avoidance subscale also showed an alpha of .94, the Food Avoidance subscale of .88, and the Intimacy and Body Exposure Avoidance subscale of .82.

### Associations with other variables

The GIAS total score demonstrated strong convergent validity through moderate to strong associations with all tested theoretically related constructs, in the expected directions (Table 3). At the subscale level, only the Food Avoidance subscale did not show significant correlations (with two variables: illness behaviour avoidance and psychological flexibility). Overall, the GIAS seems to be especially associated with illness shame, GI symptoms, and psychological distress.

**Table 3.** Correlations of the GIAS’s total score and subscales with other measures (N = 102).

|  | GIAS |  |  |  |
| --- | --- | --- | --- | --- |
|  | Total Score | General Avoidance | Food Avoidance | Intimacy and Body Exposure Avoidance |
| Illness behaviour avoidance (IBAS) | .35*** | .36*** | .11 | .36*** |
| Psychological flexibility (CompACT) | -.26*** | -.25* | -.10 | -.27** |
| Illness shame (CISS) | .79*** | .76*** | .55*** | .55*** |
| Self-compassion (SCS-SF) | -.35*** | -.30** | -.30** | -.28** |
| GI symptoms | .50*** | .50*** | .33*** | .30** |
| Anxiety (DASS-21) | .49*** | .48*** | .28** | .41*** |
| Depressive symptoms (DASS-21) | .41*** | .38*** | .20* | .41*** |
| Quality of life (EUROHIS-QOL-8) | -.35*** | -.31** | -.24* | -.32*** |

*Note.* \* $p < .05$ ; \*\* $p < .01$ ; \*\*\* $p < .001$ .

### Incremental validity

Linear regression analyses were conducted to examine the associations of GIAS and IBAS (general illness behaviour avoidance) with GI symptoms, anxiety, depressive symptoms, and quality of life (Table 4). IBAS was significantly associated with anxiety, depressive symptoms, and lower quality of life, but not with GI symptoms. GIAS was associated with all variables.

**Table 4.** Linear regression testing IBAS and GIAS as independent variables in four different models (N = 102).

|  | GI symptoms |  | Anxiety<br>(DASS-21) |  | Depressive symptoms<br>(DASS-21) |  | Quality of life<br>(EUROHIS-QOL-8) |  |
| --- | --- | --- | --- | --- | --- | --- | --- | --- |
| | $\beta$ | $R^2$ | $\beta$ | $R^2$ | $\beta$ | $R^2$ | $\beta$ | $R^2$ |
| IBAS | .14<br>(p = .114) | .25 | .23<br>(p = .013) | .27 | .36<br>(p < .001) | .27 | -.29<br>(p = .003) | .18 |
| GIAS<br>(total score) | .45<br>(p < .001) |  | .41<br>(p < .001) |  | .28<br>(p = .003) |  | -.25<br>(p = .012) |  |

Notably, GIAS contributed unique explanatory power beyond IBAS regarding GI symptoms and anxiety (although not regarding depressive symptoms and quality of life). This suggests that GIAS captures distinct variance in both GI symptom severity and anxiety levels in individuals with GI conditions.

### GIAS as mediator

A mediation analysis was conducted using PROCESS macro, with GI symptoms as independent variable, GIAS as mediator, and psychological distress as dependent variable (Figure 1). The total association (considering direct + indirect effects) of GI symptoms and psychological distress was b = .37, SE = .07, p < .001. When controlling for GIAS, this association remained significant, although at a lower magnitude (b = .23, SE = .08, p = .006). The indirect association of GI symptoms and psychological distress via GIAS was also significant (b = .14, 95% CI [.05, .26]), supporting the presence of a partial mediation effect. The total model explained 29% of the variance in psychological distress.

**Figure 1.**
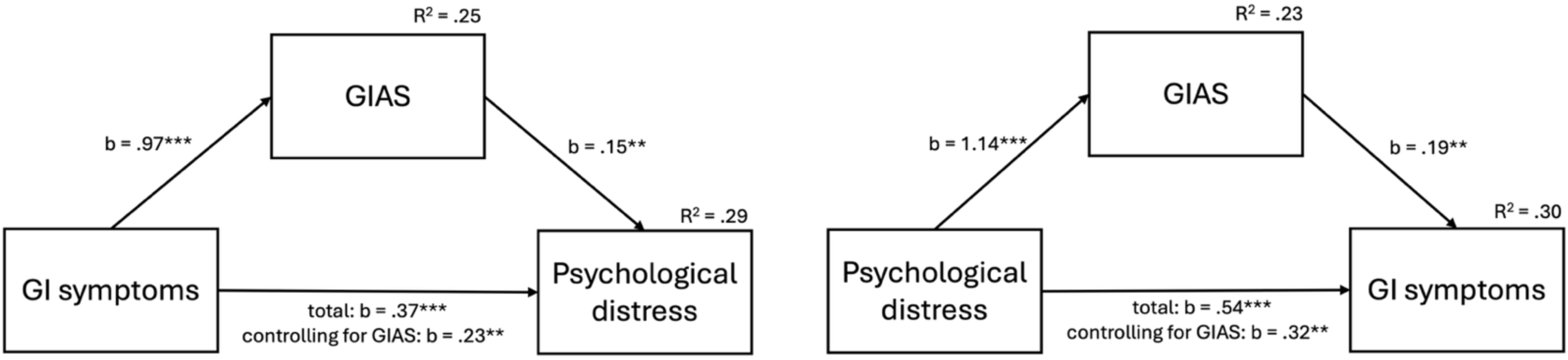
Mediation models illustrating the mediating role of GI-specific avoidance (GIAS) in the bi-directional associations of GI symptoms and psychological distress. Unstandardized coefficients (b) are shown. **p < .01; ***p < .001.

A similar analysis was then performed, with psychological distress as independent variable, GIAS as mediator, and GI symptoms as dependent variable (Figure 1). The total association of psychological distress with GI symptoms was significant (b = .54, SE = .11, p < .001). When controlling for GIAS, this association decreased in magnitude (b = .32, SE = .11, p = .006). The indirect association of psychological distress and GI symptoms, as mediated by the mechanisms captured by the GIAS, was significant (b = .21, 95% CI [.09, .39]), establishing the GIAS as a partial mediator of this association. This model accounted for 30% of the variance in GI symptoms.

## Discussion

This study presents the development and validation of GIAS, a self-report measure designed to assess patterns of avoidance relevant to individuals living with GI conditions. Drawing on empirical literature, clinical theory, and multidisciplinary expertise, the GIAS addresses an important gap in the assessment of experiential and behavioural avoidance in individuals with GI conditions, capturing three distinct and clinically meaningful domains: general avoidance, food avoidance, and intimacy and body exposure avoidance.

The factor analytic structure of the GIAS supports the theoretical and empirical assumption that GI-related avoidance is multifaceted. The emergence of a general avoidance factor reflects broader tendencies to restrict daily functioning, emotional processing, and cognitive engagement in response to GI symptoms, patterns that align with core tenets of experiential avoidance and psychological inflexibility (1, 33).

The food avoidance subscale captures the strategies of dietary restriction and avoidance of food-related anxiety/fear commonly seen across IBS, IBD, and celiac disease populations (4, 7, 9, 13, 34). Such avoidance behaviors may resemble those observed in Avoidant/Restrictive Food Intake Disorder (ARFID), where individuals limit food intake due to fear of adverse consequences, leading to nutritional deficiencies and psychosocial impairment (35). In individuals with GI conditions, food avoidance can exacerbate symptom severity and reduce quality of life, underscoring the need for targeted interventions (7). It is noteworthy to highlight that the GIAS, or its Food subscale, are not measures or screeners for ARFID but rather assess broader patterns of avoidance in response to GI symptoms, that may overlap with, but are conceptually and diagnostically distinct from, eating disorders.

The third factor, intimacy and body exposure avoidance, highlights how shame, fear of symptom visibility, and loss of control influence decisions around sexual intimacy, clothing, and physical vulnerability. These domains are often overlooked in research and clinical practice, yet central to patient quality of life, as body-related shame and concerns about symptom disclosure can lead to social withdrawal and decreased relationship satisfaction among individuals with GI disorders (19, 36, 37).

The GIAS demonstrated excellent internal consistency and strong convergent validity, with significant associations with established indicators of psychological distress, symptom severity, illness-related shame, and general illness behaviour avoidance. Importantly, the scale also showed expected negative correlations with psychological flexibility, self-compassion, and quality of life, further supporting its construct validity. The specificity of the food avoidance factor, which showed limited associations with psychological distress but retained associations with symptom severity and shame, suggests the presence of a more behaviourally delineated but clinically relevant dimension of avoidance. These findings may align with previous literature indicating that food-related fear and restriction are not always indicative of emotional distress but may nonetheless have a significant impact on functioning (12).

Incremental validity analyses showed that the GIAS explained unique variance in GI symptoms and anxiety above and beyond the IBAS. This suggests that GI-specific avoidance, as measured by the GIAS, captures clinically relevant processes that are not accounted for by more general measures of illness-related avoidance. These findings have important implications for both assessment and intervention, particularly in the context of ACT- and exposure-based therapies that explicitly target experiential avoidance (38, 39).

Moreover, mediation models in our study provided preliminary evidence that GI-specific avoidance may function as a key process between the known bidirectional link of GI symptoms and psychological distress (40). While the cross-sectional nature of the data precludes causal conclusions, these results are consistent with psychological models of chronic illness that position avoidance as both a response to symptom burden and a maintaining factor in the cycle of distress and disability (1–3).

Several limitations to this study should be acknowledged. First, the cross-sectional design prevents any conclusions about causality or temporal precedence between avoidance, symptoms, and psychological outcomes. Longitudinal and experimental designs are needed to test the scale’s sensitivity to change, and clarify the interplay between symptoms, GI-specific avoidance, and psychological distress. Further, while the sample included individuals with a range of GI diagnoses, recruitment through social media and patient associations may have led to a selection bias toward individuals more engaged, distressed, or with higher education levels, potentially limiting generalisability. Finally, while preliminary validity was established, further psychometric testing (e.g., confirmatory factor analysis, test-retest reliability, measurement invariance across diagnostic groups), including samples from different cultures and languages, is needed to establish the GIAS as a robust clinical and research instrument.

Overall, the GIAS represents a novel and transdiagnostic contribution to the assessment of psycho-behavioural processes in GI conditions. This scale is theoretically and empirically grounded, and psychometrically robust, measuring of GI-specific avoidance in its diverse forms and life domains. The GIAS can potentially support both clinical formulation and the evaluation of treatment mechanisms in interventions for individuals living with GI conditions.

## Data Availability

All data produced in the present study are available upon reasonable request to the authors.

## Acknowledgments

We thank Axel Josefsson MD PhD, Armando Peixoto PhD, and the patients that were involved in this study.

## Author contributions

Inês A. Trindade, PhD (Conceptualization: Lead; Formal analysis: Lead; Funding acquisition: Lead; Methodology: Lead; Project administration: Lead; Supervision: Lead; Validation: Equal; Writing – original draft: Lead; Writing – review & editing: Equal) Ana Pereira (Data curation: Equal; Formal analysis: Supporting; Investigation: Equal; Methodology: Equal; Writing – review & editing: Supporting)

Bruna Veloso (Data curation: Equal; Formal analysis: Supporting; Investigation: Equal; Writing – review & editing: Supporting)

Tom van Gils (Conceptualization: Supporting; Methodology: Supporting; Validation: Supporting; Writing – review & editing: Lead)

Sanna Nybacka (Conceptualization: Equal; Methodology: Supporting; Validation: Supporting; Writing – review & editing: Lead)

