## SupplementaryMaterial1-4 for "The GI-specific Avoidance Scale (GIAS): Development, psychometric validation, and incremental power of a new questionnaire"

**Supplementary Material 1**

**Full description of Procedures**

This study was approved by the Ethics and Deontology Committee of the Faculty of Psychology and Educational Sciences at the University of Coimbra (CEDI/FPCEUC: 89/7) and was conducted in accordance with the ethical standards outlined in the Declaration of Helsinki. Data collection took place in Portugal, with participants recruited through targeted outreach via social media platforms (e.g., Facebook) and national patient associations, including the Portuguese Association for Inflammatory Bowel Disease and the Crohn's Colitis Portugal Association. Eligibility criteria were: a) age 18 years or older; b) a self-reported medical diagnosis of a gastrointestinal condition (e.g., irritable bowel syndrome, inflammatory bowel disease, functional dyspepsia, celiac disease, colon cancer, or stomach cancer); c) fluency in Portuguese; and d) not being pregnant.

Participants provided informed consent electronically before initiating the survey. Data were collected between August 1 and September 30, 2024, using self-report questionnaires hosted on LimeSurvey (version 6). To ensure data quality, attention-check questions were embedded throughout the survey, following best practices for online data validation (Shamon & Berning, 2020).

Of the 255 individuals who accessed the survey, 153 did not complete it and were excluded from the final sample. The remaining 102 participants completed the entire protocol and were included in the analyses.

**Supplementary Material 2**

**Full description of Measures**

***Illness behavior avoidance (IBAS)***

The IBAS (Trindade et al., 2021) is a 7-item self-report scale designed to assess illness-related experiential avoidance. Items reflect strategies such as avoiding thoughts about the illness, steering clear of symptom-triggering situations, attempting to control symptoms, and concealing symptoms from others (e.g., "I find it difficult to talk to my family/friends/partner about my symptoms"). Responses are rated on a 5-point Likert scale ranging from 0 ("Never") to 4 ("Always").

***Psychological flexibility (CompACT-8)***

The CompACT-8 (Francis et al., 2016; Portuguese version: Trindade et al., 2021; Tyndall et al., 2023) is an 8-item version of the original 23-item Comprehensive Assessment of ACT Processes, validated for Portuguese illness groups. It measures psychological flexibility through two core subscales: Behavioral Awareness (e.g., "Even when doing the things that matter to me, I find myself doing them without paying attention") and Valued Action (e.g., "I behave in line with my personal values"). Participants rate each item on a 7-point Likert scale from 0 ("Never true") to 6 ("Always true").

***Illness shame (CISS)***

The CISS (Trindade et al., 2017) is a 7-item scale that evaluates the extent of shame experienced in relation to living with a chronic illness or its symptoms (e.g., "I feel that my illness is embarrassing"). Items are rated on a 5-point Likert scale from 0 ("Never true") to 4 ("Always true").

***Self-compassion (SCS-SF)***

The SSCS-SF(SCS-SF; Neff, 2023; Raes et al., 2011; Portuguese version: Castilho et al., 2015) is a 12-item version of the original Self-Compassion Scale. It assesses six components of self-compassion: self-kindness, self-judgment, common humanity, isolation, mindfulness, and over-identification. Items are rated on a Likert scale, from 1 ("Almost Always") to 5 ("Almost Never").

***GI symptoms***

The frequency of GI symptoms over the past month was assessed using a self-report inventory adapted from previous research. Items evaluate the occurrence of common gastrointestinal symptoms, including abdominal pain, bloating, loose stools, urgency, excessive gas, tenesmus, frequent bowel movements, nausea, and difficulty sleeping due to GI discomfort. Respondents rate each symptom on a 7-point Likert scale ranging from 0 (never) to 6 (always), with higher scores indicating higher GI symptom frequency.

***Depressive symptoms and anxiety (DASS-21)***

The DASS-21 (Lovibond & Lovibond, 1995; Portuguese version: Pais-Ribeiro et al., 2004) assesses symptoms of depression, anxiety, and stress. Each of the three subscales contains 7 items rated on a 4-point Likert scale ranging from 0 ("Did not apply to me at all") to 3 ("Applied to me very much or most of the time"), based on experiences of the previous week. In this study, only the depression and anxiety subscales were used.

***Quality of Life (EUROHIS-QOL-8)***

The EUROHIS-QOL-8 (Power, 2003; Schmidt et al., 2006; Portuguese version: Pereira et al., 2011) is an 8-item measure adapted from the WHOQOL-BREF. It assesses overall quality of life across four core domains: physical health (e.g., “Do you have enough energy for everyday life?”), psychological well-being (e.g., “How satisfied are you with yourself?”), social relationships (e.g., “How satisfied are you with your personal relationships?”), and environmental conditions (e.g., “How satisfied are you with the conditions of your living place?”).

**Supplementary Material 3**

**The 37 initial item pool of the GIAS**

1. I try not to think about my GI condition so that I don't feel anxious or depressed.
2. I rarely try new foods because I'm afraid of having GI symptoms.
3. I delay seeking medical help when my GI symptoms get worse.
4. I avoid using public toilets in fear of making noise due to my GI symptoms.
5. I avoid physical exercise due to fear of triggering GI symptoms.
6. I avoid paying attention to my GI symptoms.
7. I avoid searching for information (e.g. internet) about my GI condition.
8. Controlling my GI symptoms is one of the most important things in my life.
9. I avoid drinks or foods that I enjoy because I fear they will trigger GI symptoms.
10. I avoid going to social events for fear of showing GI symptoms.
11. I avoid talking about my GI condition, even though I would like to discuss it more.
12. I try to keep myself busy to avoid having negative thoughts or feelings about my GI symptoms.
13. When I have GI symptoms, I immediately try to hide them from other people.
14. When I have GI symptoms, I usually change my plans for that day.
15. I avoid meeting new people because I'm afraid of being judged for my GI condition.
16. I tell myself that I shouldn't have certain thoughts about my GI condition.
17. I distance myself from friends and family when my GI symptoms worsen.
18. I don't have sex as much as I would like to because I don't want to show signs of my GI condition.
19. I always try to carry medication for my GI condition with me, just in case.
20. I avoid going to restaurants because of my GI condition.
21. I don't make long-term plans due to the unpredictability of my GI condition.
22. I avoid leaving home when I believe other will notice my GI symptoms.
23. Even though sometimes I would like to try new foods, I usually don't because I don't know how they would affect my GI symptoms.
24. I avoid wearing clothes I like, especially tight ones, due to my GI symptoms (for example, feeling bloated).
25. I only make plans to go out when I know there will be a toilet nearby.

26. I would be a much more active person if I didn’t have to control my GI symptoms all the time.
27. I avoid going to long social events (e.g. weddings, baptism ceremonies, birthday parties, work gatherings) because of my GI condition.
28. I don’t have sex as much as I would like to due to my GI symptoms.
29. I avoid being naked in front of others because I don’t want to show signs of my GI condition.
30. I avoid certain foods because I fear they will worsen my GI symptoms.
31. The way I feel I have to eat due to my GI condition has limited my lifestyle.
32. I avoid having sex because I’m afraid of faecal incontinence or passing gas.
33. I avoid using public transports due to my GI symptoms.
34. I make an effort to avoid thinking about my fear of having a new flare related to my GI condition.
35. I try not to be too far from a toilet to avoid feeling anxious.
36. I struggle attending social events because I’m afraid of having GI symptoms in public.
37. I struggle telling people I have this GI condition.

*Response scale:*

0 - Strongly disagree

1 - Mostly disagree

2 - Slightly disagree

3 - Slightly agree

4 - Mostly agree

5 - Strongly agree

**Supplementary Material 4**

**The Portuguese (validated) version of the final 20 items of the GIAS**

*Instruções:*

Por favor, leia cada uma das afirmações abaixo e indique o grau de concordância, de 0 a 5, de cada um dos seguintes itens, usando a escala:

0 - Discordo totalmente
1 - Discordo bastante
2 - Discordo ligeiramente
3 - Concordo ligeiramente
4 - Concordo bastante
5 - Concordo totalmente

12. Tento manter-me ocupado/a para impedir que surjam pensamentos ou sentimentos negativos sobre os meus sintomas gastrointestinais. [1]

14. Quando tenho sintomas gastrointestinais, costumo mudar os meus planos para esse dia. [1]

15. Evito conhecer pessoas novas com medo de ser julgado/a pela minha condição gastrointestinal. [1]

17. Tendo a afastar-me de amigos e família quando os meus sintomas gastrointestinais se agravam. [1]

21. Não faço planos a longo prazo devido à imprevisibilidade da minha condição gastrointestinal. [1]

25. Planeio sair de casa apenas quando sei que haverá uma casa de banho próxima. [1]

33. Evito usar transportes públicos devido aos meus sintomas gastrointestinais. [1]

34. Esforço-me para não pensar no medo de uma nova crise relacionada com a minha condição gastrointestinal. [1]

35. Tento não estar muito longe de uma casa de banho para evitar sentir-me ansioso/a. [1]

36. Tenho dificuldade em ir a eventos sociais porque tenho medo de ter sintomas gastrointestinais em público. [1]

2. Raramente experimento novas comidas porque tenho medo de ter sintomas gastrointestinais. [2]

9. Evito bebidas ou comidas de que gosto porque tenho medo que causem sintomas gastrointestinais. [2]

20. Evito ir a restaurantes por causa da minha condição gastrointestinal. [2]

30. Evito comer algumas comidas por medo de agravar os meus sintomas gastrointestinais. [2]

31. A forma como sinto que tenho de comer devido à minha condição gastrointestinal restringiu muito o meu estilo de vida. [2]

18. Não tenho tantas relações sexuais como gostaria porque não quero mostrar sinais da minha condição gastrointestinal. [3]

24. Evito usar roupas de que gosto (por exemplo, justas ao corpo), por causa dos meus sintomas gastrointestinais (por exemplo, sentir-me inchado/a). [3]

29. Evito estar nu/nua à frente de outras pessoas porque não quero mostrar sinais da minha condição gastrointestinal. [3]

32. Evito ter relações sexuais com medo de ter incontinência fecal ou gases. [3]

--

*Legend:*

[1] General Avoidance subscale

[2] Food Avoidance subscale

[3] Intimacy and Body Exposure Avoidance subscale
